# Automated epidural spinal cord stimulation for cardiovascular regulation in spinal cord injury: from optimization to real-time implementation

**DOI:** 10.64898/2026.07.21.26358253

**Authors:** Breanne Christie, Siqi Wang, Harley Ledbetter, Lauren Diaz, Harrison Nguyen, Gail F. Forrest, Nathan Torgerson, Claudia A. Angeli, Erik C. Johnson, Susan J. Harkema, Francesco V. Tenore

## Abstract

**Background:** Spinal cord injury (SCI) is frequently associated with orthostatic hypotension, defined by a sustained decrease in blood pressure upon assuming an upright posture due to impaired autonomic regulation. Cardiovascular spinal cord epidural stimulation (CV-scES) can regulate systolic blood pressure (SBP) in people with SCI, but stimulation paradigms are highly individualized. To make this treatment available to more patients, we developed an algorithm to tailor individualized CV-scES paradigms that closely mimic researcher-developed paradigms.

**Methods:** We performed an offline analysis using datasets collected from eight individuals with SCI with epidural stimulators implanted over the lumbosacral spinal segments. During data collection, researchers modulated stimulation parameters with the goal of maintaining SBP between 110-120 mmHg. Each two-hour dataset included synchronized SBP and stimulation recordings. We ran optimization analyses offline to determine temporal requirements before modifying stimulation amplitude to mitigate out-of-range SBP.

**Results:** The algorithm parameters that best matched researcher-selected stimulation changed relatively quickly during the first 12 min (one every ∼40 sec), and more slowly thereafter (one every ∼79 sec). Overall, algorithmic stimulation closely tracked researcher-controlled stimulation, with a mean correlation coefficient of 0.94. To evaluate online performance, we tested the algorithm in real time with a single participant. We found that a faster approach was needed to respond to changes in SBP caused by rapid, unpredictable events, such as postural changes. We implemented a sigmoid-based paradigm that determined the time to wait before changing stimulation as a function of the current SBP, with worse SBP values requiring faster responses. The new paradigm outperformed the original algorithm and researcher-controlled stimulation across measures of SBP stability, though recovery from a postural tilt maneuver remained slower than with researcher control.

**Conclusions:** Our results indicate that algorithmic stimulation may minimize assistance required from researchers and participants, making CV-scES more feasible for clinical translation.

## 1 Background

Spinal cord injury (SCI) can not only impair sensory and motor function, but it can also have a profound impact on the autonomic nervous system. This often manifests as blood pressure instability, such as orthostatic hypotension and autonomic dysreflexia. The rates of orthostatic hypotension immediately after injury can reach as high as 82% for people with tetraplegia and 50% for people with paraplegia [1] and can persist for many years [2]–[4]. Orthostatic hypotension results from impaired regulation of blood pressure during postural changes [5]. Blood pressure typically decreases when moving from lying to standing and increases when returning to a supine position; these limitations impose strict limits on the ability of people with SCI to participate in rehabilitation [5]. Chronic hypotension can further result in reduced cognitive function [6]–[9], decreased quality of life [10], and increased risk of stroke [11], and can be life threatening [6], yet there are few viable treatment options. Previous studies of people with SCI have found that chronic hypotension can be managed by a combination of patient education and the use of pharmacologic and nonpharmacologic interventions, however blood pressure remains very unstable even with these approaches [5], [12]. Compression garments can mitigate hypotension because they aid venous return, but they are burdensome to use in daily life [13]. Pharmacological methods can increase mean arterial pressure for people with SCI, but they do not stabilize blood pressure, are controversial in addressing the cognitive deficits of orthostatic hypotension, and may exacerbate episodes of autonomic dysreflexia [14]–[17]. People with SCI typically adapt to blood pressure instability without expectation of resolution, leading to diminished quality of life [5], [18].

A newer approach, cardiovascular targeted spinal cord epidural stimulation (CV-scES), can stabilize blood pressure in people with chronic SCI [19]–[26]. Prior studies delivered CV-scES via an electrode array implanted at T11-L1 vertebral levels over spinal cord segments L1-S1. When delivering CV-scES during postural stress, orthostatic hypotension was ameliorated, indicating that CV-scES can improve cardiovascular regulatory systems [21]. When CV-scES was terminated, participants’ blood pressures returned to baseline values within 15 mins [20]. Once the investigators found an effective open-loop/tonic stimulation paradigm in the laboratory setting, the participants could use the device at home for up to six hours per day. For some individuals, baseline blood pressure actually improved after using CV-scES at home for a few months [21].

Despite the vast potential of CV-scES, stimulation paradigms are highly individualized. To make this treatment option accessible to a larger population, it is critical to automate the paradigm development. One approach for doing this is to use learning algorithms to facilitate more efficient and effective stimulation paradigms. Due to the vast number of potential stimulation combinations (e.g., stimulation polarity, pulse amplitude, pulse width, and pulse frequency across each electrode), it is not feasible for a study team to evaluate every possible paradigm. This approach also relies on training researchers to identify complex patterns in multimodal data. Identifying trends and automating this process has the potential to improve how responsive participants are to CV-scES and could save time, while potentially providing additional insights into the underlying mechanisms of how CV-scES works.

Therefore, our goal for this study was to develop an algorithm that produces individualized stimulation paradigms that closely mimic those implemented by research investigators. We aimed to minimize the difference between algorithm-derived and investigator-selected stimulation amplitude over time while maintaining systolic blood pressure within a clinically-defined target range. This approach was designed to avoid overfitting and improve generalizability to real-time, closed-loop implementation. In the long term, the algorithm is intended to support clinical implementation of closed-loop stimulation while incorporating user input.

## 2 Methods

### 2.1 Research participants

Data were collected from eight people with cervical-level SCI (Table 1). All of the participants presented with orthostatic hypotension, persistent chronic low blood pressure when at rest, and routine symptoms of autonomic dysreflexia with no cardiovascular disease unrelated to SCI. Each individual was previously implanted with an array of 16 electrodes (5-6-5 Specify; Medtronic, Minneapolis, MN, USA) over lumbosacral spinal cord segments. The array was subcutaneously connected to a pulse generator (Intellis™; Medtronic) implanted in the flank [27], [28].

**Table 1:** Research participant characteristics. AIS = American Spinal Injury Association Impairment Scale. M=male, F=female.

| Subject ID | Sex | Time since injury (years) | Spinal level of injury | AIS grade<sup>576</sup> |
| --- | --- | --- | --- | --- |
| A64 | M | 38 | C4 | A<sup>578</sup> |
| A97 | M | 15 | C4 | A |
| A119 | F | 11 | C5 | A<sup>579</sup> |
| A128 | M | 9 | C4 | A |
| B38 | M | 7 | C4 | B<sup>580</sup> |
| B42 | M | 7 | C4 | B |
| C193 | F | 9 | C4 | B<sup>581</sup> |

### 2.2 Training data for offline analysis

We utilized two offline experimental datasets from participant A64, three from A97, three from A119, one from A128, three from B38, eight from B42, three from C193, and five from C237. The datasets were collected anywhere from 41-589 days after device implantation, and were selected because they involved similar experimental protocols. Each dataset contained ∼2 hours of CV-scES, with ∼15 mins of rest/baseline time before and after CV-scES. The goal of the experiments was to maintain systolic blood pressure between 110 and 120 mmHg, a healthy range determined by the American Heart Association [29].

During the experiments, the participants were seated upright in their wheelchairs and were asked to minimize movements. The participants were asked to verbally report when they felt a muscle spasm, which was noted by experimenters. Adjustments to the stimulation paradigm (e.g., pulse amplitude, frequency, width) were denoted by a synchronized pulse to identify the event on the acquired data. The beat-to-beat systolic and diastolic blood pressure and heart rate were recorded every ∼0.9 sec using photoplethysmography of the index finger, middle finger, or thumb (Finapres Medical Systems, Amsterdam, Netherlands). To ensure the photoplethysmography measurements were accurate, they were compared to readings from a sphygmomanometer several times throughout each experiment and recalibrated when needed.

### 2.3 Algorithm design for offline analysis

The primary focus of the initial algorithm was on modulating stimulation amplitude over time in response to changes in SBP, such that low SBP values triggered an increase in amplitude and high SBP triggered a decrease, with the step size defined as the median change in amplitude calculated from the corresponding offline dataset. The algorithm’s starting stimulation amplitude value was the same that the researchers utilized. All other stimulation parameters remained fixed during this testing period.

We performed an optimization analysis to identify the optimal latency before implementing a change in stimulation amplitude when SBP was out of the clinically-defined target range (110-120 mmHg). In the first iteration of the algorithm (v1), to account for noise in the natural fluctuation of SBP, the SBP had to remain out of range for a minimum of 5 sec before modifying amplitude. Using a grid search optimization approach, we evaluated latencies from 5-180 sec to identify which values produced the smallest difference between algorithmically-derived amplitude and expert-driven amplitude, deemed to be the “error.” The amplitude step size and the latency range were selected based on discussions with the investigators. They were the main variables that the researchers tracked when developing stimulation paradigms, so for the first pass of the algorithm, our goal was to recreate that capability. In the future, we will incorporate other stimulation parameters to make the algorithm more advanced and surpass what is possible with human-designed stimulation paradigms.

After observing that the investigators changed stimulation amplitude more frequently or in larger step sizes during the beginning of a session, we interpreted this as an initial broad search for optimal parameters. As the parameter space narrows and blood pressure approaches the target range, adjustments become more incremental. This motivated our implementation of two distinct temporal phases in the algorithm. To determine which phase lengths would result in the lowest error relative to expert-driven amplitude changes in the second iteration of the algorithm (v2), we set the duration of the first phase to 4-110 mins of stimulation; we evaluated 1-minute intervals from 4-20 mins and in 5-min intervals from 25-110 mins. Once the optimal phase length across datasets was determined, we again used a grid search approach to identify four optimized latency parameters for each dataset: (1) the time to wait before decreasing stimulation in phase 1, (2) the time to wait before decreasing stimulation in phase 2, (3) the time to wait before increasing stimulation in phase 1, and (4) the time to wait before increasing stimulation in phase 2. To characterize the similarities between algorithmic and expert-selected stimulation amplitudes, we ran a linear regression between them for each dataset with both the single-phase (v1 of the algorithm) and the dual-phase scenarios (v2 of the algorithm).

### 2.4 Real-time algorithm design and testing

Following the offline analysis, we conducted a real-time implementation test with Participant A64 consisting of three phases: researcher-controlled amplitude tuning, implementation of v1 of the algorithm, and implementation of a revised version of the algorithm (v3, see details below). As in the original recordings, the participant was seated upright and blood pressure was recorded every 0.9 sec. In this paradigm, algorithmic recommendations for amplitude adjustments were generated in real-time in an app on a tablet, which were manually approved by the researcher before being input for delivery. Baseline data were collected for the first 10 mins, during which time persistent hypotension was observed. Researchers then used a 30 min period to manually adjust stimulation amplitude to bring the SBP into the target range of 110-120 mmHg. After 20 mins of stimulation, the participant’s chair was tilted backwards to approximately 115° (25° posterior to upright) for approximately five minutes, altering gravitational loading relative to the fully upright seated position. This positional change was expected to increase SBP due to SCI-related hemodynamic instability and changes in the relationship between the implant lead and the spinal cord. The resulting increase in SBP necessitated a reduction in the amount of delivered current.

An additional 10 mins of baseline data without stimulation were then collected, followed by 30 mins of single-phase algorithm (v1) deployment. This initial version was selected as a baseline implementation because it represents the simplest control strategy, allowing us to evaluate whether a single-phase approach could adequately regulate SBP before introducing more complex adaptations. Based on what the researchers implemented in the most recent experimental session, we set the amplitude step size to the median value of 0.3 mA, time to wait before decreasing stimulation to 92 sec, and time to wait before altering stimulation to 75 sec. Stimulation amplitudes were recommended by the v1 algorithm and implemented by a researcher to ensure participant safety. After stimulation was delivered for 18 mins, the participant underwent another tilt maneuver for five minutes.

Another 10 mins of baseline data without stimulation were then collected, and a new version of the algorithm (v3) was subsequently deployed for 30 mins. While the dual-phase approach (v2) improved agreement with investigator-selected parameters relative to the single-phase approach (v1), it relied on fixed latency values that limited responsiveness to rapid changes in SBP, such as those induced by postural perturbations. In v3, we replaced the fixed waiting-time parameters with a sigmoid-based paradigm (*Equations 1 and 2*) that determined the time to wait before changing stimulation as a function of the current SBP. When SBP exited the target range, the algorithm initiated a waiting period whose duration decreased as SBP deviated further from the target range and was canceled if SBP returned to range before a stimulation adjustment occurred. For Equation 1, which corresponded to increasing amplitude in response to low SBP, T_min_ = 6 sec, T_max_ = 75 sec (matches the parameter found in v1), k = 0.3, x_m_ = 82.5, and Δ = 0.8. For Equation 2, which corresponded to decreasing amplitude in response to high SBP, T_min_ = 27 sec, T_max_ = 92 sec (matches v1 parameter), k = 0.3, x_m_ = 147.5, and Δ = 0.8. As in the previous iterations, amplitude was adjusted in the appropriate direction by the median amplitude change used by the researcher, which was 0.3 mA. This approach preserved the core structure of the single-phase algorithm (v1) while removing the need to explicitly optimize discrete waiting times. After the v3 algorithm had been tested for 18 mins, the participant underwent another tilt maneuver for five minutes.

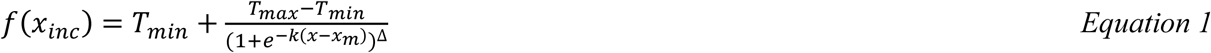

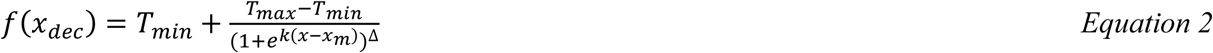

To characterize how effectively SBP was restored to the target range, we quantified the proportion of time SBP remained within that range during the pre-tilt period and across the full interval spanning both the pre-tilt and tilt perturbation periods. We also calculated the time it took for SBP to return to the target range after the tilt was initiated.

To assess SBP stability, we calculated the standard deviation and two additional outcome measures previously validated against blood pressure recordings obtained during cardiovascular perturbation protocols and extended monitoring sessions in individuals both with and without SCI [30]. Outcomes included (i) total deviation from the midpoint of the target range (115 mmHg) and (ii) a summary metric derived from the cumulative distribution of SBP values relative to the target range. Total deviation was defined using the 5th and 95th percentiles of the SBP distribution to limit the influence of outliers, and calculated as the sum of the positive differences between the 95th percentile and 115 mmHg and between 115 mmHg and the 5th percentile. The cumulative distribution curve was generated by iteratively expanding the target range in 5 mmHg increments, with lower and upper bounds constrained to 40 mmHg and 230 mmHg, respectively. At each expansion step, the percentage of SBP measurements within the expanded range was calculated. The x-axis was mapped linearly from 0 to 1 across expansion steps, and the area under the curve (AUC) was computed using trapezoidal integration. An AUC value of 100% indicates that all SBP measurements were within the target range [14].

## 3 Results

### 3.1 Offline algorithm testing

We observed that increases in stimulation amplitude generally resulted in increases in SBP and decreases in amplitude led to decreases in SBP. Each offline dataset contained between 1-55 amplitude adjustments, and the median amplitude change was between 0.2-1.2 mA across all datasets (Supplemental Table 1).

When treating the two hour experiment as a single phase in the first iteration of the algorithm (v1), we found that the optimal time to wait before decreasing amplitude in response to high SBP was 70 ± 9 sec (mean ± standard error across all datasets). The optimal time to wait before increasing amplitude in response to low SBP was 65 ± 9 sec (Supplemental Table 1). A linear regression between the algorithmic and expert-driven amplitude resulted in a mean Pearson correlation coefficient of 0.85. When we divided each experiment into two phases in the second iteration of the algorithm (v2), we found that setting the length of phase 1 to 12 mins resulted in the highest algorithm performance, though all lengths of phase 1 less than 65 mins resulted in a higher performance than the single-phase algorithm (Figure 1). With a phase 1 length of 12 mins, the optimal time to wait before decreasing amplitude was 34 ± 6 sec during phase 1 and 68 ± 9 sec during phase 2 (Supplemental Table 2). The optimal time to wait before increasing amplitude was 45 ± 7 sec during phase 1 and 89 ± 10 sec during phase 2. In 22/28 datasets, the optimal time to wait before decreasing amplitude was shorter in phase 1 than in phase 2. Similarly, the optimal time to wait before increasing amplitude was shorter in phase 1 than in phase 2 for 21/28 datasets. On average, across all participants and datasets, the optimal time to wait before increasing or decreasing amplitude was 39.5 sec in phase 1 and 78.5 sec in phase 2. The stimulation paradigm produced by the dual-phase algorithm better replicated the researcher-driven amplitude than did the single-phase algorithm, with the mean correlation coefficient increasing to 0.94. Representative datasets are shown in Figure 2.

**Figure 1:**
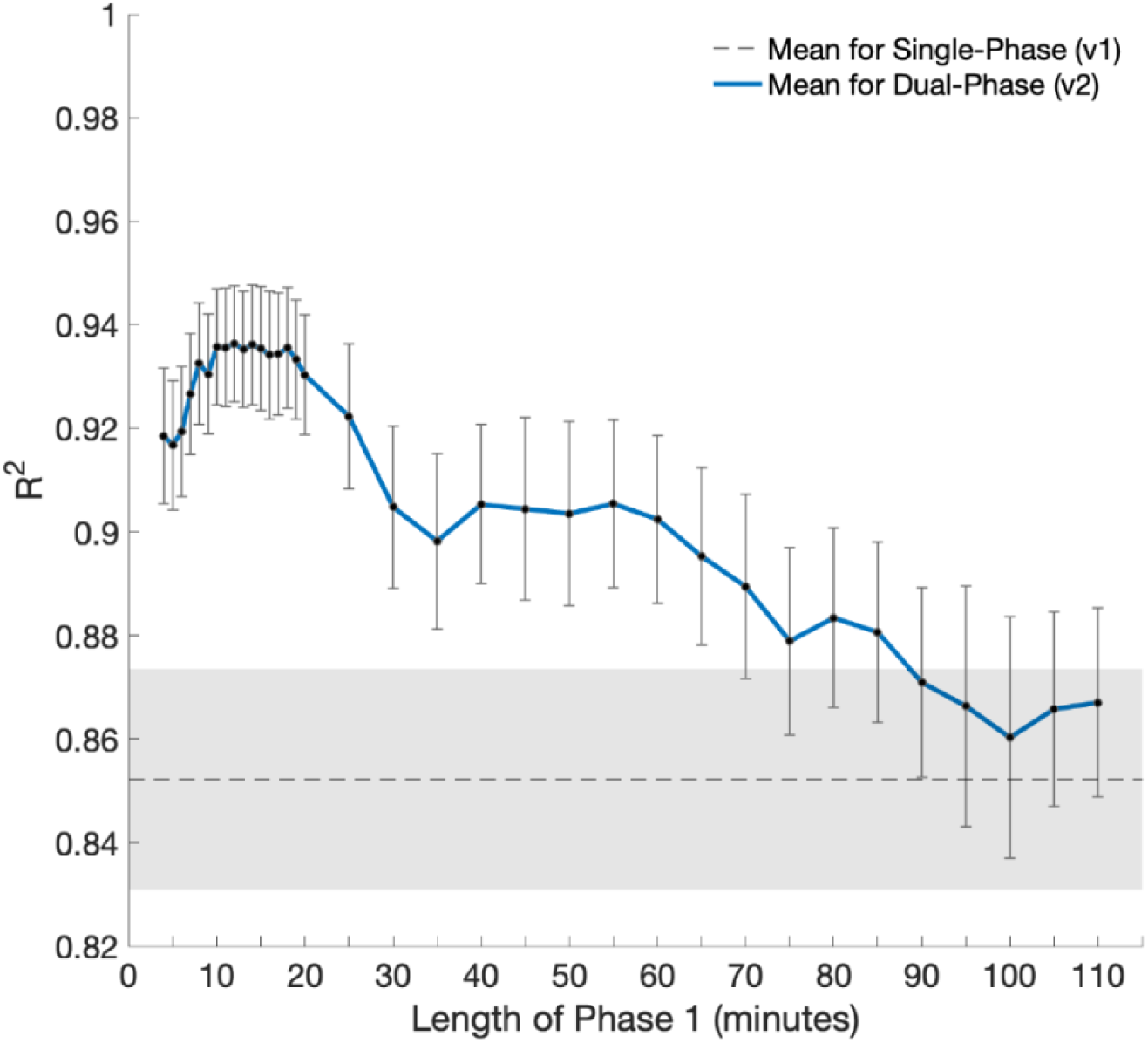
Optimization results for finding the optimal length of the first phase of CV-scES for v2 of the algorithm. Standard error is represented by the bars for dual-phase data and the shaded area around the dashed line for the single-phase data.

**Figure 2:**
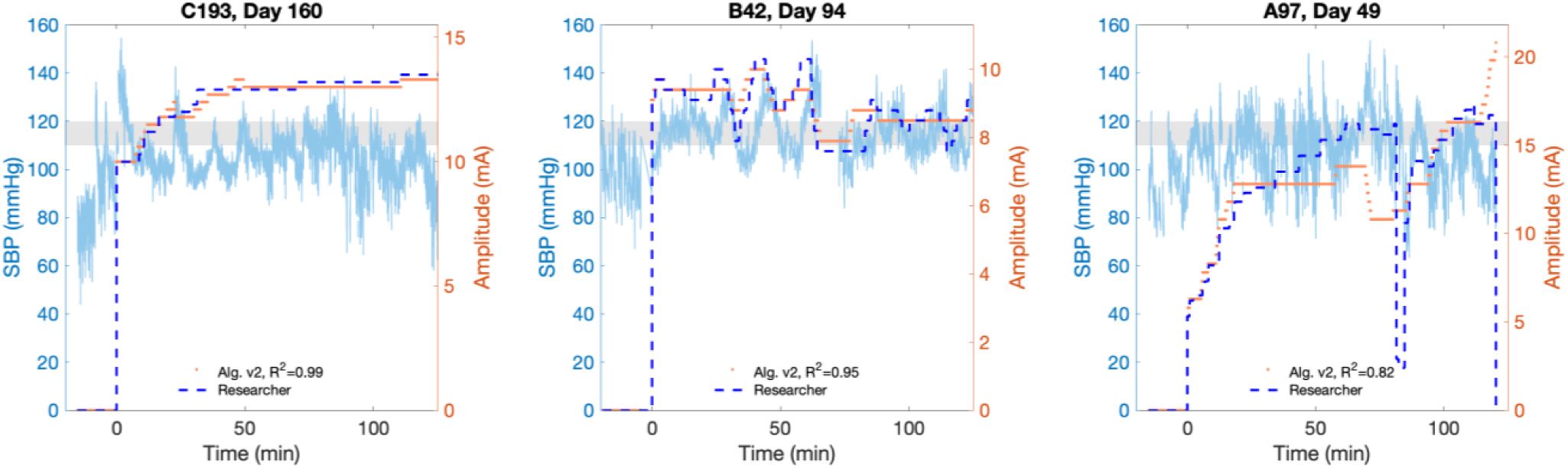
Representative results from three participants after finding the optimal time to wait in each temporal phase (length of phase 1 = 12 mins) of v2 of the algorithm before increasing or decreasing stimulation amplitude in response to low or high SBP, respectively. Systolic blood pressure is on the left y-axis and stimulation amplitude is on the right y-axis. Time = 0 represents when CV-scES began. The shaded grey region represents the target SBP range. The left plot represents an ideal scenario for the dual-phase algorithm (v2): many amplitude adjustments occurred early in the experiment, followed by steady-state behavior during the middle to end of the experiment. The middle plot represents a scenario in which the single-phase algorithm (v1) would have been sufficient, as frequent changes to stimulation amplitude were made throughout the experiment, but the dual-phase algorithm still performed well. The right plot represents a scenario in which human-reported spasm data would be critical: a muscle spasm occurred around minute 80, temporarily elevating SBP and requiring the researchers to quickly reduce the stimulation level.

### 3.2 Real-time algorithm testing

When the researchers manually tuned stimulation, amplitude was changed 23 times. Amplitude was changed 15 times when v1 of the algorithm was implemented and 10 times when v3 was tested. The temporal evolution of SBP and amplitude during each experiment phase is shown in the top row of Figure 3.

**Figure 3:**
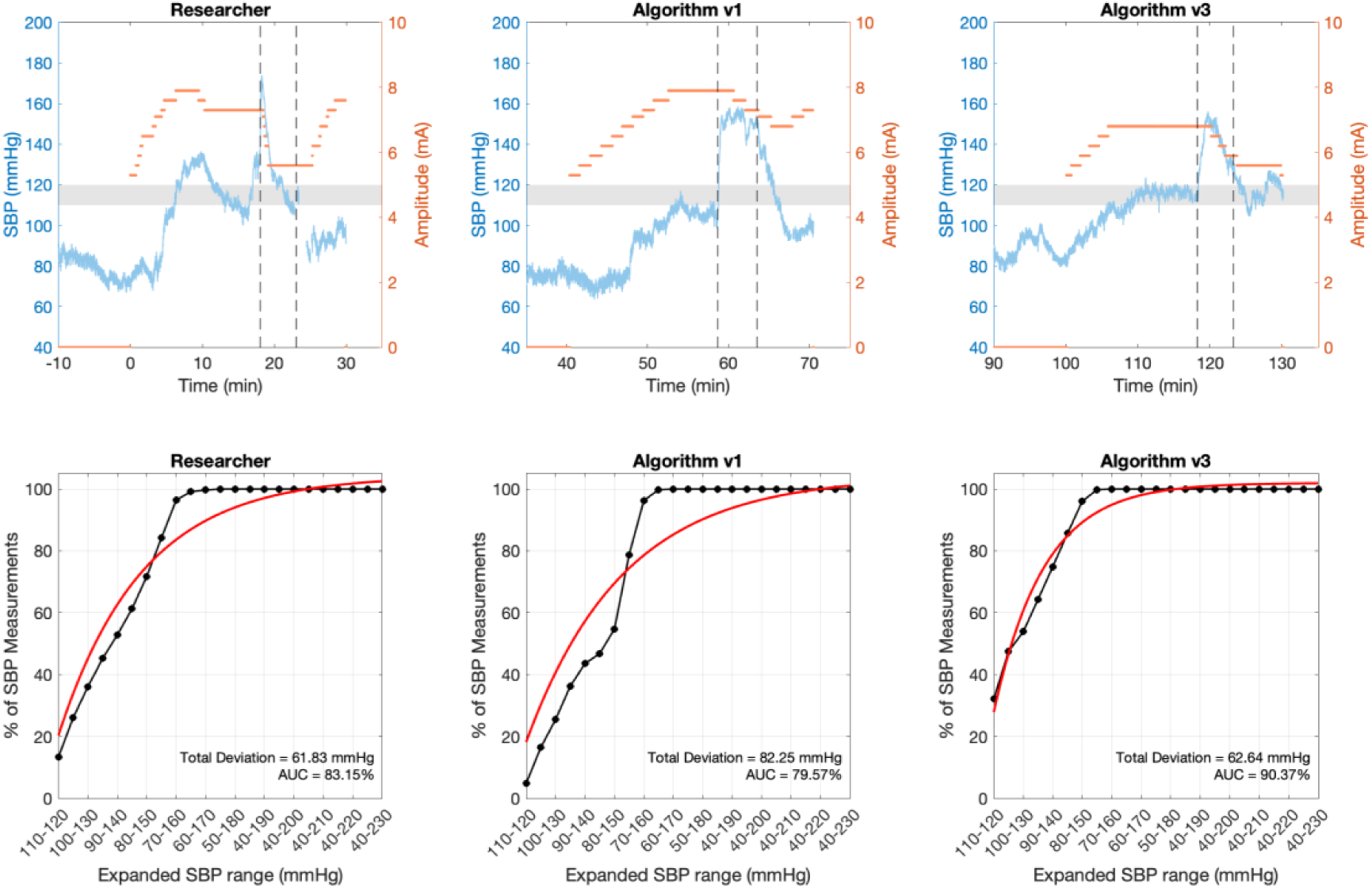
Top row: SBP traces during researcher-controlled stimulation (left), algorithm v1 (middle), and algorithm v3 (right). Dashed vertical lines indicate the tilt maneuver, during which time SBP rapidly increased. Researcher-controlled stimulation involved frequent parameter adjustments and overshoot prior to stabilization. Algorithm v1 produced slower corrective responses and limited time within the target range. Algorithm v3 more rapidly restored SBP to the target range and maintained improved stability prior to tilt while requiring fewer stimulation adjustments than manual control. Bottom row: cumulative SBP distribution curves for each condition. Black markers indicate the empirical cumulative distribution of SBP measurements across progressively expanded ranges centered on the target SBP range (110-120 mmHg), and the red line indicates the fitted exponential curve. Curves that rise more rapidly and saturate at smaller range expansions reflect tighter clustering of SBP values around the target range and improved SBP stability. Total deviation from the midpoint of the target range (115 mmHg) and AUC metrics are shown for each condition.

We observed that during the three baseline periods, the SBP never reached the target range of 110-120 mmHg. When the researchers manually tuned stimulation, SBP was in the target range 20.4% of the time before the tilt was administered (Figure 4). SBP stayed within range just 6.3% of the time when v1 of the algorithm was tested, and 52.4% of the time when v3 of the algorithm was tested. When including the tilt, SBP stayed within range 19.7% of the time for the researchers, 6.4% for algorithm v1, and 43.0% for algorithm v3. As shown in the top row of Figure 3, researcher-controlled stimulation resulted in rapid parameter adjustments with overshoot prior to stabilization, v1 exhibited slower corrective responses and prolonged excursions outside the target range, and v3 more quickly brought SBP into the target range and better maintained stability prior to tilt.

**Figure 4:**
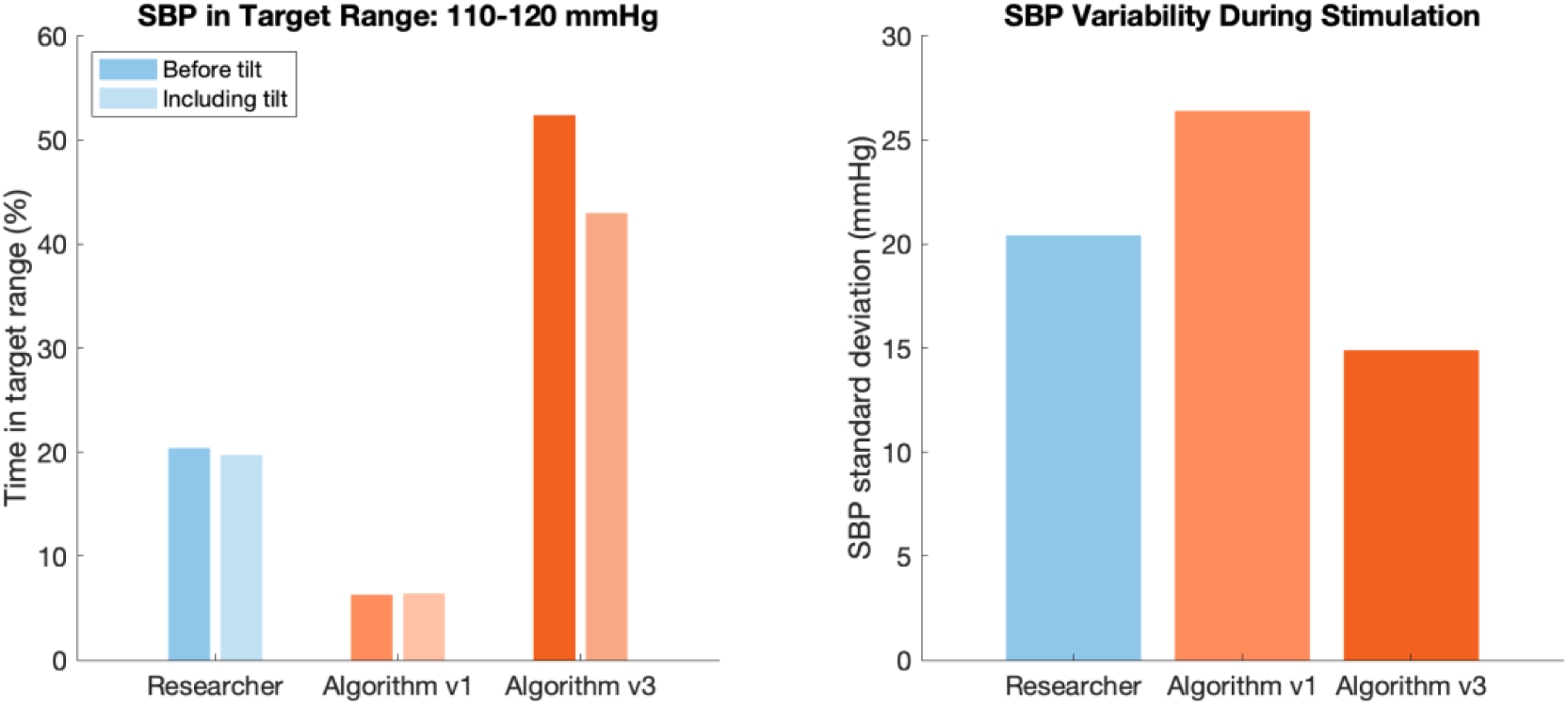
Left: Percentage of time SBP remained within the target range (110-120 mmHg) before tilt and across the full stimulation period including tilt for researcher-controlled stimulation, v1 of the algorithm, and v3 of the algorithm. v3 maintained SBP within the target range substantially longer than both researcher-controlled stimulation and v1. Right: Standard deviation of SBP during stimulation for each condition. v3 produced the lowest SBP variability, indicating improved blood pressure stability relative to researcher-controlled stimulation and v1.

With respect to SBP stability, the standard deviation across the period of stimulation was 20.42 mmHg when the researchers manually tuned stimulation parameters, 26.40 mmHg when v1 of the algorithm was tested, and 14.90 mmHg when v3 of the algorithm was tested (Figure 4). The total deviation was 59.58 mmHg for the researchers, 83.44 mmHg for v1, and 57.16 mmHg for v3 (Figure 3). In comparison, a previous study found that 13 control subjects typically had a total deviation of 32.5 ± 4.9 mmHg (mean ± standard error) over a period of 24 hours [31]. The AUC was 88.75% when the researchers manually tuned stimulation parameters, 82.35% for v1, and 93.69% for v3 (Figure 3).

In all three tilt maneuvers, the participant’s SBP rose immediately to above 150 mmHg, requiring a decrease in delivered current. The researchers returned SBP to the target range more quickly than either version of the algorithm, requiring 2.1 mins compared to 6.1 mins for v1 and 4.5 mins for v3.

## 4 Discussion

The goal of this study was to develop robust spinal cord epidural stimulation algorithms that closely mimic individualized paradigms developed by research investigators. Our results demonstrate that algorithmic control can approximate expert-driven stimulation both offline and in real time, improving performance of stabilizing blood pressure relative to an initial algorithm version and, in some metrics, approaching or exceeding manual tuning by researchers. These findings support the feasibility of using automated approaches to regulate blood pressure while reducing reliance on continuous expert adjustment.

In accordance with prior studies, we observed that increasing current (by increasing stimulation amplitude) resulted in increases in SBP, and decreasing current led to decreases in SBP towards the starting baseline value [19]–[21]. The experts that collected these data, who are authors of this study, typically changed the amplitude in small intervals (≤1.2 mA) when attempting to reach the optimal SBP range between 110-120 mmHg. The step sizes were defined in part by hardware limitations of the device. They were also done in part for the comfort of the participant and to avoid unintended skeletal muscle activity of the lower extremities. As each participant’s trends were characterized across multiple experimental sessions, the stimulation paradigms became more refined and required less broad parameter exploration within a session. Early sessions involved a wider search over stimulation parameters to identify an effective range, whereas later sessions began closer to an established operating point and required only minor adjustments. As a result, datasets collected at different stages of this process exhibited variability in stimulation parameters, and consequently algorithm parameters, within the same participant. In later sessions, the distinction between exploratory and fine-tuning phases was often reduced, as less optimization was required overall. Future algorithm development would focus on the individualized optimization and expanding to other stimulation parameter predictions.

The algorithm better fit the data when we divided each dataset into two phases, with the optimal length of the first phase being the first 12 mins of stimulation. Though any length of time ≤65 mins allowed the algorithm to predict the stimulation amplitude better than when treating stimulation as a single phase, shorter times are preferable in order to reach a steady stimulation state more quickly. Our motivation for implementing these phases was indeed based on observing data and noticing that the investigators made more frequent changes to the stimulation paradigm in the first half of each experiment. This behavior was adopted because the first half of the experiment was typically used to identify optimal stimulation including intensity, while the second half was used to steadily maintain SBP within the target range. Additionally, we observed that the research team tended to wait less time to decrease stimulation amplitude in response to high SBP than they waited before increasing amplitude for low values. This asymmetry likely reflects the clinical priority of avoiding hypertensive events.

Importantly, real-time testing demonstrated that iterative refinement of the algorithm translated into meaningful improvements in closed-loop SBP regulation. While the initial real-time implementation (v1) spent limited time within the target range and responded slowly to perturbations, the revised version (v3) more rapidly brought SBP into range, maintained improved stability prior to tilt, and was associated with fewer stimulation adjustments than manual tuning. Although recovery following tilt remained faster under researcher control, v3 of the algorithm reduced overshoot and achieved greater overall time within the target range. These findings suggest that adjusting the waiting time before making stimulation changes according to SBP deviation may improve responsiveness while maintaining SBP stability. Incorporating position, absolute blood pressure, and level of muscle activation into this control strategy may further support autonomous blood pressure regulation.

The present study was indeed limited by a small sample size; in future work, we hope to expand the study to include additional participants and expand to additional stimulation parameters. A larger sample size will enable us to identify trends between algorithm parameters and participant characteristics, which we will use to establish more generalizable stimulation paradigms. Next, we will incorporate stimulation parameters such as pulse frequency, pulse width, and cathode/anode selection into the algorithm. We will also include a “human in the loop” feature that allows a user to provide additional contextual information, such as the occurrence of muscle spasms or use of the spinal cord stimulator for other functions, including walking [32] or voiding the bladder [33].

## 5 Conclusions

This study made advances towards making CV-scES a more accessible approach for improving cardiovascular function. For people with SCI, restoring cardiovascular function and limiting abnormalities in blood pressure can increase independence and improve quality of life [10]. Our results indicate that algorithmic stimulation can approximate expert-driven stimulation both offline and in real time. Relative to the initial algorithm version, the updated algorithm improved blood pressure stability and, in some metrics, approached or exceeded manual tuning by researchers. These findings support the feasibility of using automated approaches to regulate blood pressure while reducing reliance on continuous expert adjustment.

## Data Availability

The datasets used and/or analyzed during the current study are available from the corresponding author upon reasonable request.

## 6 List of abbreviations

AIS: American Spinal Injury Association Impairment Scale
AUC: Area under the curve
CV-scES: Cardiovascular spinal cord epidural stimulation
IRB: Institutional Review Board
SCI: Spinal cord injury
SBP: Systolic blood pressure
scES: Spinal cord epidural stimulation
SE: Standard error
Tmax: Maximum wait time
Tmin: Minimum wait time

## 7 Declarations

### 7.1 Ethics approval and consent to participate

The study protocol was approved by both the University of Louisville and Kessler Foundation Institutional Review Boards. Informed consent was obtained prior to participation in research-related activities.

### 7.5 Authors’ contributions

BC, SW, GF, CA, EJ, SH, and FT contributed to conception and design of the study. HL conducted the experiments and SW processed the experimental data. BC wrote the first draft of the manuscript. All authors contributed to manuscript revisions, and read and approved the submitted version.

## 7.6 Acknowledgements

The authors would like to thank our research participants for their courage, dedication, motivation, and perseverance that made these research findings possible. We also thank the entire research staff for administrative, engineering, surgical, medical, and additional research support.

---

**Supplemental Table 1:** Parameters that result in the highest algorithm performance for v1 of the algorithm. Day # = number of days since surgery.

| Subject ID | Day # | Median amplitude change (mA) | Time (sec) to wait before decreasing amplitude | Time (sec) to wait before increasing amplitude |
| --- | --- | --- | --- | --- |
| A64 | 44 | 0.3 | 88 | 9 |
|  | 48 | 0.3 | 61 | 18 |
| A97 | 49 | 0.5 | 10 | 17 |
|  | 58 | 0.6 | 64 | 29 |
|  | 76 | 0.2 | 17 | 39 |
| A119 | 121 | 0.6 | 38 | 97 |
|  | 134 | 0.9 | 5 | 58 |
|  | 188 | 0.9 | 41 | 19 |
| A128 | 56 | 0.5 | 35 | 19 |
| B38 | 512 | 1.2 | 36 | 36 |
|  | 552 | 0.9 | 44 | 70 |
|  | 589 | 0.9 | 16 | 74 |
| B42 | 94 | 0.3 | 69 | 72 |
|  | 133 | 0.6 | 180 | 36 |
|  | 195 | 0.9 | 135 | 60 |
|  | 211 | 0.6 | 117 | 54 |
|  | 240 | 0.6 | 140 | 64 |
|  | 267 | 0.6 | 76 | 41 |
|  | 293 | 0.4 | 94 | 51 |
|  | 295 | 0.6 | 64 | 40 |
| C193 | 41 | 0.85 | 69 | 24 |
|  | 55 | 0.9 | 32 | 75 |
|  | 160 | 0.3 | 109 | 142 |
| C237 | 93 | 0.3 | 107 | 156 |
|  | 163 | 0.3 | 69 | 142 |
|  | 215 | 0.6 | 38 | 115 |
|  | 309 | 0.9 | 162 | 86 |
|  | 344 | 0.9 | 33 | 180 |
| Mean ± std error | | 0.62 ± 0.05 | 70 ± 9 | 65 ± 9 |

**Supplemental Table 2:** Parameters that result in the highest algorithm performance for v2 of the algorithm. Phase 1 = 0-12 mins of CV-scES. Phase 2 = 12-120 mins of CV-scES. Day # = number of days since surgery.

| Subject ID | Day # | Time (sec) to wait before decreasing amplitude in phase 1 | Time (sec) to wait before decreasing amplitude in phase 2 | Time (sec) to wait before increasing amplitude in phase 1 | Time (sec) to wait before increasing amplitude in phase 2 |
| --- | --- | --- | --- | --- | --- |
| A64 | 44 | 30 | 84 | 6 | 25 |
|  | 48 | 103 | 92 | 6 | 121 |
| A97 | 49 | 5 | 21 | 16 | 24 |
|  | 58 | 64 | 70 | 45 | 27 |
|  | 76 | 5 | 27 | 42 | 39 |
| A119 | 121 | 5 | 38 | 82 | 121 |
|  | 134 | 5 | 5 | 68 | 58 |
|  | 188 | 41 | 22 | 10 | 27 |
| A128 | 56 | 27 | 55 | 15 | 37 |
| B38 | 512 | 5 | 34 | 28 | 43 |
|  | 552 | 5 | 44 | 42 | 98 |
|  | 589 | 12 | 17 | 46 | 89 |
| B42 | 94 | 23 | 58 | 62 | 45 |
|  | 133 | 70 | 180 | 35 | 38 |
|  | 195 | 11 | 119 | 59 | 53 |
|  | 211 | 25 | 159 | 20 | 127 |
|  | 240 | 20 | 56 | 30 | 69 |
|  | 267 | 48 | 85 | 16 | 164 |
|  | 293 | 37 | 109 | 19 | 100 |
|  | 295 | 13 | 133 | 21 | 111 |
| C193 | 41 | 69 | 17 | 16 | 27 |
|  | 55 | 30 | 54 | 44 | 180 |
|  | 160 | 109 | 11 | 45 | 160 |
| C237 | 93 | 36 | 107 | 27 | 119 |
|  | 163 | 15 | 69 | 109 | 142 |
|  | 215 | 38 | 30 | 104 | 100 |
|  | 309 | 82 | 166 | 65 | 180 |
|  | 344 | 5 | 33 | 180 | 180 |
| Mean ± std error | | 34 ± 6 | 68 ± 9 | 45 ± 7 | 89 ± 10 |

